# Assessment of *LIN28A* variants in Parkinson’s disease

**DOI:** 10.1101/2020.06.10.20103291

**Authors:** Monica Diez-Fairen, Mary B. Makarious, Sara Bandres-Ciga, Cornelis Blauwendraat, on behalf of the International Parkinson’s Disease Genomics Consortium (IPDGC)

## Abstract

Parkinson’s disease (PD) is a complex neurodegenerative disease with a strong genetic component in which both rare and common genetic variants contribute to disease risk, onset and progression. Despite that several genes have been associated with familial forms of disease, validation of novel genes associated with PD remains extremely challenging. Recently, a heterozygous loss-of-function variant in LIN28A was associated with PD pathogenesis in the Asian population. Here, we comprehensively assess the role of LIN28A variants in PD susceptibility using individual-level genotyping data from 14,671 PD cases and 17,667 controls, as well as whole-genome sequencing data from 1,647 PD patients and 1,050 controls. Additionally, we further assessed the summary statistics from the most recent GWAS meta-analyses to date for PD risk and age at onset. After evaluating these data, we did not find evidence to support a role for LIN28A as a major causal gene for PD. However, additional large-scale familial and case-control studies in non-European ancestry populations are necessary to further evaluate the role of LIN28A in PD etiology.

## Correspondence

Parkinson’s disease (PD) is a complex neurodegenerative disorder with a strong genetic component in which both rare and common genetic variants contribute to disease risk, onset and progression. Mutations in several genes have been associated with familial forms of disease, most of which are highly penetrant rare variants resulting in early onset presentation. Although significant progress in understanding the genetic basis of PD has been made, validation of novel genes associated with PD remains extremely challenging (Blauwendraat *et al*, 2020; Bandres-Ciga *et al*, 2020).

We read with great interest the recently published article by *Chang and colleagues* in which the authors report and claim that a rare variant in *LIN28A*, which encodes for a highly conserved RNA-binding protein (Lin28a), contributes to PD pathogenesis (Chang *et al*, 2019). Through whole exome sequencing, the authors identified a heterozygous missense variant in *LIN28A* (p.Arg192Gly, rs558060339), shown to result in gene loss-of-function (LOF), in two patients of East Asian ancestry with early-onset PD (EOPD). In addition, midbrain dopamine (mDA) neurons differentiated from patient-derived human embryonic stem cells showed developmental defects and PD-specific pathological features that could be rescued by expression of wild-type Lin28a. Indeed, previous work from the same group had shown that loss of *LIN28A* function could ultimately lead to a PD-like phenotype coupled with mDA neuronal loss (Rhee *et al*, 2016).

Here, we sought to investigate the role of *LIN28A* variants in PD susceptibility as part of the International Parkinson’s Disease Genomics Consortium (IPDGC) efforts to screen reported risk factors for PD. We used genome-wide association study (GWAS) data comprising 14,671 PD cases and 17,667 neurologically healthy individuals, and publicly available whole-genome sequencing (WGS) data including 1,647 PD patients without known disease-causing mutations (mean age at onset (AAO) 64.2 ± 9.6), of which 145 cases had EOPD (mean AAO 45.2 ± 4.6), and 1,050 controls (mean age 60.3 ± 11.9) from the Accelerating Medicines Partnership - Parkinson’s disease initiative (AMP-PD; www.amp-pd.org). Additionally, we further assessed data from the most recent GWAS meta-analyses to date (excluding 23andMe data) for PD risk consisting of 15,056 PD cases, 18,618 UK Biobank proxy-cases (i.e., subjects with a first degree relative with PD) and 449,056 controls (Nalls *et al*, 2019), as well as for age at onset comprising 17,996 PD cases (Blauwendraat *et al*, 2019). All participants were of European ancestry and quality control procedures on both individual and variant levels are described in detail elsewhere (Nalls *et al*, 2019; Blauwendraat *et al*, 2019).

We identified a total of 83 variants within the *LIN28A* gene in WGS data, of which only three were coding, including two synonymous (p.Lys127Lys, p.Pro205Pro) and one non-synonymous variant (p.Thr189Ile) (**Table 1**). All three coding variants are extremely rare (MAF ≤ 1%) according to public databases. Case-control association testing using single-variant score test adjusted by covariates (including sex, age, family history, education level, 10 principal components, and cohort) showed no significant differences in the frequency of *LIN28A* variants between PD cases and controls after Bonferroni correction (significance threshold = 0.05/83 = 6.02E-04) (**Table 1, Supplementary Table 1**). Gene-based burden analyses did not detect an enrichment of rare variants in PD cases versus controls (**Supplementary Table 2**).

**Table 1.**
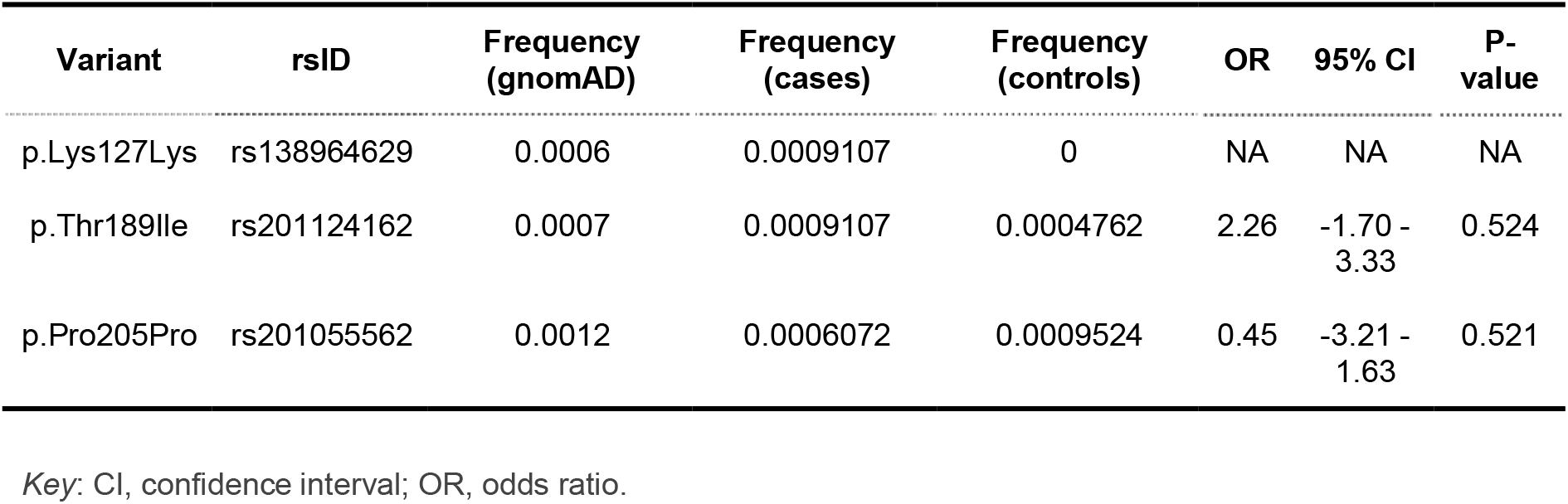
*LIN28A* coding variants in whole-genome sequencing data from PD and controls.

Similarly, we performed a case-control association analysis using logistic regression in imputed individual-level genotyping data from IPDGC. All 34 *LIN28A* variants found were non-coding, including 28 intronic and six in the 3’-untranslated region (3’-UTR). None of the variants were significantly associated with the disease (**Supplementary Table 3**). We further analyzed summary statistics from the above-mentioned risk and age at onset PD GWAS meta-analyses. No evidence for an association between *LIN28A* common genetic variation and PD was detected for either PD risk or age at onset (**Supplementary Figure 1; Supplementary Table 4**). Among the 97 *LIN28A* variants identified within summary statistics, 81 were intronic, 14 were in the 3’-UTR, and only two were coding non-synonymous variants (p.Arg123Gln, p.Thr189Ile) (**Supplementary Table 4**). A total of 46 variants (47.4%) were rare (MAF ≤ 3%). Of note, we did not find the original reported mutation (NC_000001.10:g.26752893C>G, p.Arg192Gly) by *Chang and colleagues* in any of our datasets.

Furthermore, we explored the frequency of LOF variants in *LIN28A* by looking at the gnomAD database v.2.1.1 (https://gnomad.broadinstitute.org/), which comprises a total of 125,748 exomes and 15,708 genomes. Six LOF variants were identified (four stop-gain and two frameshift, all heterozygous) resulting in an estimate of 0.0056% of the population having haploinsufficiency for *LIN28A*. The reported p.R192G variant was identified in six Korean individuals out of a total of 1909 included, setting the allele frequency at 0.15% in the general population.

In conclusion, based on the current data presented, our analyses do not support a role for *LIN28A* variants as a major causal gene or risk factor for PD. However, the vast majority of our data is from European ancestry, in which the p.Arg192Gly variant is not present according to the gnomAD database, and we have a relatively small number of EOPD cases in the cohorts used. Noteworthy the frequency of this variant is relatively high in the Korean population. Overall, we cannot rule out a potential role of very rare variants in *LIN28A* in PD among other populations.

Additional large-scale familial and case-control studies in non-European ancestry populations are necessary to further evaluate the role of *LIN28A* in PD etiology.

## Data Availability

The data that support the findings of this study are avail- able from the corresponding author, upon reasonable request.

## Acknowledgements

We would like to thank all of the subjects who donated their time and biological samples to be a part of this study. We also would like to thank all members of the International Parkinson’s Disease Genomics Consortium (IPDGC). For a complete overview of members, acknowledgements and funding, please see http://pdgenetics.org/partners. This work was supported in part by the Intramural Research Programs of the National Institute of Neurological Disorders and Stroke (NINDS), the National Institute on Aging (NIA), and the National Institute of Environmental Health Sciences both part of the National Institutes of Health, Department of Health and Human Services; project numbers 1ZIA-NS003154, Z01-AG000949-02 and Z01-ES101986. In addition, this work was supported by the Department of Defense (award W81XWH-09-2-0128), and The Michael J Fox Foundation for Parkinson’s Research. Data used in the preparation of this article were obtained from the AMP PD Knowledge Platform. For up-to-date information on the study, visit https://www.amp-pd.org. AMP PD – a public-private partnership – is managed by the FNIH and funded by Celgene, GSK, the Michael J. Fox Foundation for Parkinson’s Research, the National Institute of Neurological Disorders and Stroke, Pfizer, and Verily. We would like to thank AMP-PD for the publicly available whole-genome sequencing data, including cohorts from the Fox Investigation for New Discovery of Biomarkers (BioFIND), the Parkinson’s Progression Markers Initiative (PPMI), and the Parkinson’s Disease Biomarkers Program (PDBP). This work utilized the computational resources of the NIH HPC Biowulf cluster (http://hpc.nih.gov).

